# Neutralising antibodies predict protection from severe COVID-19

**DOI:** 10.1101/2022.06.09.22275942

**Authors:** Deborah Cromer, Megan Steain, Arnold Reynaldi, Timothy E. Schlub, Sarah C. Sasson, Stephen J. Kent, David S. Khoury, Miles P. Davenport

## Abstract

**Background:** Vaccine protection from COVID-19 has been shown to decline with time-since-vaccination and against SARS-CoV-2 variants. Protection against severe COVID-19 is higher than against symptomatic infection, and also appears relatively preserved over time and against variants. Although protection from symptomatic SARS-CoV-2 infection is strongly correlated with neutralising antibody titres, this relationship has been less well described for severe COVID-19. Here we analyse whether neutralising antibody titre remains predictive of protection against severe COVID-19 in the face of waning neutralising antibody levels and emerging variants.

**Methods:** We extracted data from 15 studies reporting on protection against a range of SARS-CoV-2 clinical endpoints (‘any infection’, ‘symptomatic infection’ and ‘severe COVID-19’). We then estimated the concurrent neutralising antibody titres using existing parameters on vaccine potency, neutralising antibody decay, and loss of recognition of variants and investigated the relationship between neutralising antibody titre and vaccine effectiveness against severe COVID-19.

**Findings:** Predicted neutralising antibody titres are strongly correlated with vaccine effectiveness against symptomatic and severe COVID-19 (Spearman *ρ* = 0.94 and 0.63 respectively, *p*<.001 for both). The relationship between neutralisation titre and protection is consistent with previous estimates, with 76% (127 of 167) of reported values of protection against severe COVID-19 across a range of vaccines and variants lie within the 95% confidence intervals of the previously published model.

**Interpretation:** Neutralising antibody titres are predictive of vaccine effectiveness against severe COVID-19 including in the presence of waning immunity and viral variants.

**Funding:** National Health and Medical Research Council (Australia), Medical Research Future Fund (Australia).

**Evidence before this study**
COVID-19 vaccine effectiveness against symptomatic SARS-CoV-2 infection has been shown to be strongly predicted by neutralising antibody titres. However, there has not been sufficient data on vaccine efficacy against severe disease to determine whether the correlation between neutralising antibody titres and protection is maintained for severe COVID-19. Indeed, the apparent faster waning of vaccine efficacy against symptomatic (compared to severe) COVID-19 has led some to hypothesise about a ‘decoupling’ of protection from symptomatic and severe disease. It is therefore important to identify whether neutralising antibody titre remains a correlate of protection against severe COVID-19 in real-world scenarios of waning immunity and SARS-CoV-2 variants. We searched PubMed between inception and March 2, 2022 for studies (Key search terms: (SARS-CoV-2 OR COVID-19) AND (followup OR waning OR duration OR durable) AND (protection OR efficacy OR effectiveness)) and also monitored other public sources of information such as Twitter. We identified 15 studies that reported data on vaccine effectiveness against (i) a defined clinical endpoint (ii) for an identifiable variant, (ii) for a single vaccine (or vaccine type), (iii) over an identified time since vaccination, and (iv) for which data was either provided in or readily extractable from the original publication.

**Added value of this study**
Previous work has identified that neutralising antibodies correlate strongly with protection from symptomatic COVID-19 disease for both ancestral virus and for variants of concern. Here we examine published data on vaccine effectiveness to determine if this correlation remains valid for predicting protection against severe COVID-19. Our work shows that vaccine effectiveness against severe COVID-19 is strongly correlated with neutralising antibody titres for different vaccines, variants, and over the first six months after vaccination. The relationship between vaccine effectiveness and protection against severe COVID-19 is consistent with a previously published analysis and indicates that the 50% protective titre for protection against severe COVID-19 is lower than that associated with protection from symptomatic infection.

**Implications of all of the available evidence**
In the face of increased exposure and immunity to numerous SARS-CoV-2 variants it is becoming increasingly important to be able to predict not only protection against symptomatic infection, but also protection against severe COVID-19. Here we show that neutralising antibody titres remain predictive of vaccine effectiveness against severe COVID-19 in the face of waning immunity and SARS-CoV-2 variants. This is consistent with a low level of neutralising antibodies being associated with protection from severe COVID-19. This work will be of use in providing early estimates of protection against severe COVID-19 for new SARS-CoV-2 antigenic variants, informing optimal booster timing, and will support future vaccine development by allowing prediction of vaccine protection conferred against severe outcomes.

## Introduction

Immunisation against SARS-CoV-2 has been shown to be highly effective in preventing both mild and severe COVID-19^1-3^. Previous work has shown that neutralising antibody responses are highly predictive of vaccine efficacy against symptomatic SARS-CoV-2 infection^4-7^. However, waning of antibody titres and the emergence of SARS-CoV-2 variants with extensive immune escape from vaccine-induced neutralising antibodies has contributed to declining vaccine efficacy against symptomatic infection^8-12^. Despite the loss of vaccine efficacy against symptomatic infection, protection against hospitalisation and death has remained high, leading some to speculate on an apparent ‘decoupling’ of the mechanisms of protection against mild and severe COVID-19^13^.

Studies of the relationship between neutralising antibodies and protection from symptomatic SARS-CoV-2 infection have shown that neutralising antibody titres are highly predictive of vaccine efficacy and that a titre of ∼20% of the early convalescent level is associated with 50% protection from symptomatic infection^4^. This relationship predicted that a decline in neutralising antibody titres, either as a result of waning immunity or changes in SARS-CoV-2 viral variants, will lead to reduced vaccine protection against COVID-19^4^. Analysis of protection against SARS-CoV-2 variants suggests that neutralising antibody levels remain predictive of protection against the Alpha, Beta, Delta, and Omicron variants^14,15^. It was also demonstrated that efficacy against severe outcomes was associated with a lower neutralising antibody titre, and it was estimated that the 50% protective titre against severe COVID-19 is approximately 6.5-fold lower than against symptomatic infection. However, this finding relied on data from five phase 3 studies with a combined total of only 60 severe cases. Thus it was not possible to directly demonstrate a correlation between neutralisation titres and protection from severe outcomes (Figure 1A)^4^. Importantly, this lower level of neutralising antibodies associated with protection from severe COVID-19 (compared to mild disease) directly predicted that protection against severe infection will persist for longer as antibody levels wane and will be better maintained against new variants^4^ (Figure 1A). However, the relationship between neutralising antibody titres and protection from severe COVID-19 has not been definitively tested (largely because of the small number of severe cases observed in the phase 3 vaccine trials). In this study we aggregated data from multiple observational studies, which report on vaccine efficacy and effectiveness over time and against different SARS-CoV-2 variants, to investigate whether neutralising antibodies provide a correlate of protection against severe COVID-19.

**Figure 1.**
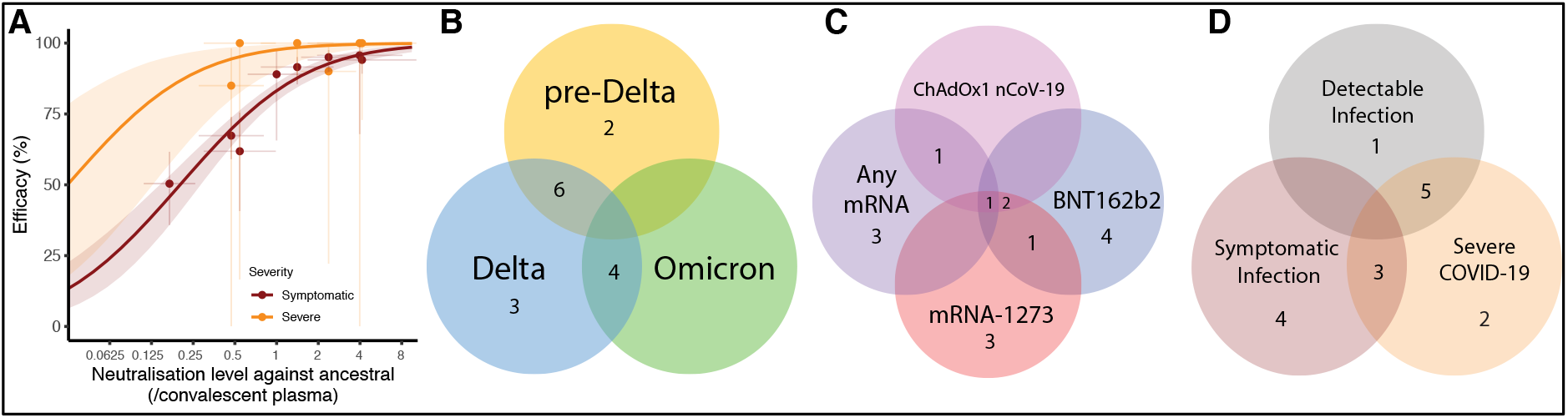
(A) Previously reported relationship between neutralising antibody titre and vaccine efficacy in prevention of symptomatic (dark red) and severe (orange) COVID-19 (reproduced from Khoury et. al.4). Solid lines indicate best fit model and shaded areas indicate 95% confidence intervals. Neutralisation titre and efficacy data used to parameterise the model is indicated as dots (95% CI indicated as whiskers). (B – D) Summary of the clinical studies used in this analysis (Table S1).

## Results

### Analysis of vaccine effectiveness in epidemiological studies

To understand vaccine effectiveness against severe SARS-CoV-2 infection, we searched the literature for studies that reported vaccine effectiveness against symptomatic and severe COVID-19 where results were reported by vaccine, circulating variant(s), and time since vaccination. We identified and extracted data from 15 studies that reported vaccine efficacy or effectiveness in this way. These were comprised of 2 randomised controlled trials, 8 test-negative case-control studies (TNCC), and 5 retrospective cohort studies (see Supplementary Materials and Table S1). This included studies of BNT162b2 (8 studies), mRNA-1273 (7 studies), ChAdOx1 nCoV-19 (4 studies) and any mRNA vaccine (that aggregated BNT162b2 and mRNA-1273 vaccines) (5 studies). These studies reported protection against pre-Delta (predominantly ancestral (Wuhan-like) and Alpha, 7 studies), Delta (13 studies) and Omicron (4 studies) variants. The studies reported vaccine protection against detectable infection (6 studies), symptomatic infection (7 studies) and severe COVID-19 (10 studies). We note that several studies reported on more than one variant or vaccine. A summary of the vaccines and variants used in the studies analysed is shown in Figure 1B-D.

Figure 2 presents the aggregated data on vaccine effectiveness in preventing severe COVID-19 for different vaccines, variants and time-since-vaccination. We used a multiple regression model to investigate the impact of vaccine type, variant, and time since vaccination on vaccine effectiveness against severe COVID-19 (Table S2). This showed that the reported effectiveness against severe COVID-19 was described best by a model that varied by vaccine, variant and over time (equation 1 and Figure 2), with the time effect dependent on variant. For example, vaccination with mRNA-1273 showed a higher effectiveness than vaccination with BNT162b2 (4.5% 95%CI 1.1 - 7.8) or ChAdOx1 nCoV-19 (8.7%, 95%CI 4.4 - 13). Similarly, effectiveness against severe COVID-19 from both the Delta and Omicron variants was lower than against the pre-Delta variants (4.7% lower, 95% CI –2.2 - 11.7 and 27.2% lower, 95%CI 18.8 - 35.5 respectively). We also found that effectiveness declined over time since vaccination, with a decrease in effectiveness of 1.2% (95% CI 0.5 - 2.0%) per month for the Delta variant and 4.4% (95%CI 3.1 - 5.6) per month for the Omicron variant. All these findings are in line with what would be expected as a result of previously reported changes in neutralising antibody levels for the different vaccines^4^, variants^14^ and time since vaccination^4,16^. We therefore next consider whether these shifts in vaccine effectiveness against severe outcomes over time for different vaccines and to variants are correlated with neutralising antibody titres.

**Figure 2.**
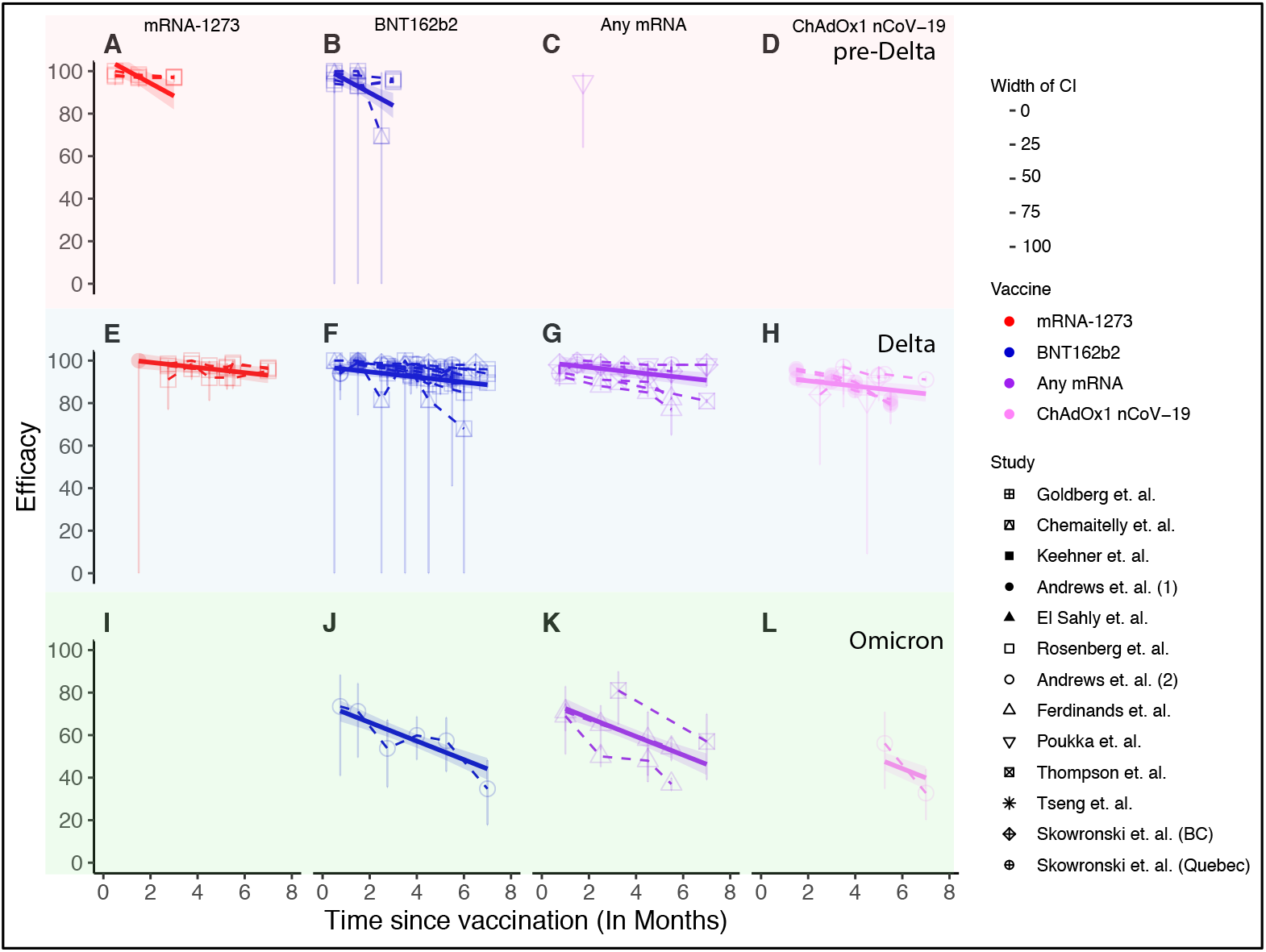
Vaccine effectiveness data against severe COVID-19 extracted from the meta-analysis (points and whiskers for 95% CI) are shown for pre-Delta (top row, panels A-D), Delta (middle row, panels E-H) and Omicron (bottom row, panels I-L) variants. Note that for panels D and I no effectiveness data was. Shading indicates the degree of confidence in the data as determined by the width of the confidence interval. Predicted vaccine effectiveness using a multiple regression model (solid lines) and 95% confidence intervals (shaded area) are overlaid.

### Correlation between neutralising antibody titre and vaccine effectiveness against severe COVID-19

To understand the relationship between *in vitro* neutralisation titre and protection we aggregated data on neutralisation titre and effectiveness across all the studies. The epidemiological studies of vaccine effectiveness did not include contemporaneous measurements of neutralising antibody titres against the different variants. However, estimates for neutralising antibody titres could be calculated for each reported real-world effectiveness value in the meta-analysis based on the vaccine, variant, and time since vaccination. That is, the expected geometric mean neutralisation titre corresponding to each vaccine effectiveness estimate can be predicted based on: (i) the geometric mean neutralisation titre (GMT) previously estimated for each vaccine^4,14,17^, (ii) the rate of waning of neutralising antibody levels^4,16^, and (iii) the drop in neutralisation titre to variants (detailed methods and parameters are given in the methods, supplementary materials, Tables S3 and S4^4^ and equations 1 and S4).

For example, Chemaitelly *et. al*.^10^ measured vaccine protection against pre-Delta and Delta variants after vaccination with the BNT162b2 vaccine. This included follow up for more than 25 weeks after vaccination, with vaccine effectiveness reported for the periods 0-4, 5-9, 10-14, 15-20, 21-25 and ≥25 weeks. The peak GMT titre for BNT162b2 vaccinees against the ancestral variant are estimated to be 2.4-fold (95% CI = 1.5-3.8)^4^ of the convalescent GMT. Neutralisation titres against the Delta variant are estimated to be 3.9-fold lower (95% CI= 3.1-4.9)^14^ than against the ancestral variant, and neutralising antibody titres have been estimated to wane with a half-life of 108 days (95% CI= 82-159)^4^. Therefore, after accounting for the initial neutralisation titre fold-drop against the variant, decay of antibodies, and the length of the original clinical trials (see supplementary methods), the GMT of BNT162b2 vaccinees against the Delta variant over the reported periods is estimated to be 0.55 (95% CI .33-.91), 0.46 (95% CI .27-.76) and 0.38 (95% CI .22-.64)-fold of the convalescent GMT against the ancestral virus for 5-9, 10-14, and 15-20 weeks post vaccination respectively (confidence intervals were obtained by bootstrapping, see supplement). The corresponding real-world vaccine effectiveness estimates of vaccine efficacy against severe COVID-19 caused by the Delta variant in these periods are 100% (95% CI 74.3-100), 81.6% (95% CI 0-99.6) and 100% (95% CI 0-100) respectively (confidence limits as reported in the clinical studies).

The estimates of neutralising antibody titre and data on vaccine effectiveness from the 15 studies included in the meta-analysis were then aggregated and compared. We observe an excellent correlation between the predicted neutralisation titre (x-axis) and reported vaccine effectiveness obtained from our meta-analysis (Spearman correlation *ρ* =.94 and 0.63 for symptomatic and severe efficacy respectively, p<.001 for both, Figure 3A and B). We explored whether this association might be driven by group differences between study types, vaccines, or variants (Supplementary Figure 1). However, the strong correlation between predicted neutralisation titre and protection from severe SARS-CoV-2 remained across these different subgroupings.

**Figure 3.**
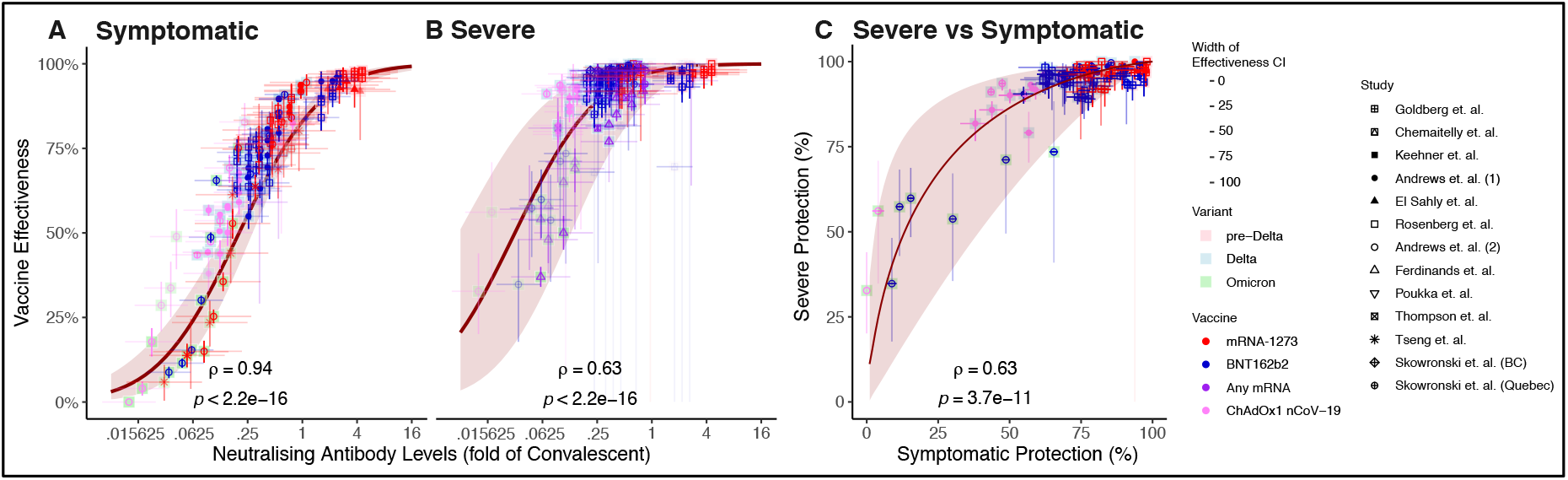
Correlation between estimated neutralising antibody titres (accounting for vaccine used, variant studied and time since vaccination) and clinical data for (A) vaccine effectiveness against symptomatic SARS-CoV-2 infection, (B) vaccine effectiveness against severe COVID-19. (C) Correlation between vaccine effectiveness against symptomatic and severe COVID-19. Solid lines indicate the predicted relationship taken from4, and shading indicates 95% CI of the model estimates. X-axis confidence intervals in A and B represent the degree of confidence in the estimate neutralising antibody titre. A breakdown of the relationship shown in panels A and B by variant and study type is shown in Figure S3.

### Neutralising antibody titres predict protection from symptomatic and severe COVID-19

The analysis above shows a strong correlation between predicted antibody titres and observed protection against acquisition of symptomatic and severe COVID-19. However, this does not provide a direct means of predicting vaccine effectiveness based on neutralisation titres. We have previously published a mathematical model relating neutralising antibody titre to vaccine protection (based on the results of the phase 3 trials for seven vaccines and protection seen in convalescent subjects against infection with the ancestral SARS-CoV-2). We therefore next plotted the reported vaccine effectiveness from our meta-analysis against the estimated neutralising antibody titres and compared this to our previously reported relationship between antibody levels and protection (Figure 3A and B). Figure 3A shows the previously reported relationship between neutralising antibody titre and protection from symptomatic SARS-CoV-2 infection (red line and shaded 95% confidence intervals, reproduced from reference^4^) overlaid with the 144 values of estimated neutralisation titre / reported effectiveness combinations we were able to obtain against symptomatic infection. Similarly, the 167 values for protection from severe COVID-19 are plotted with the predicted curves (Figure 3B). We observe that 127 / 167 (76%) of real-world estimates for vaccine efficacy against severe COVID-19 lie within the confidence intervals of the prediction. Of the estimates that lay outside the confidence intervals 29/40 (72.5%) had both reported and predicted efficacies of above 90% (i.e. they were at the very right hand side of Figure 3B and S2B where the confidence intervals are very narrow).

Predicted vaccine efficacy and reported vaccine effectiveness are also highly correlated (Pearson p-value < 0.001 for both symptomatic and severe infection. See Figure S2), consistent with the previously reported relationship between neutralisation titre and protection^4^. Similarly, we also generated the mean estimate and 95% confidence intervals for predicted vaccine effectiveness over time across different vaccines, variants and over time, for both symptomatic and severe COVID-19 (Figures S3 and S4). Despite the heterogeneity in epidemiological setting and trial design of the clinical studies, the previously reported model^4^ predicting vaccine efficacy based on neutralising antibody titres was in very good agreement with vaccine effectiveness against both symptomatic and severe COVID-19.

### Effectiveness against symptomatic infection predicts effectiveness against severe COVID-19

The analysis above shows that the previously derived relationship between neutralising antibody titres and protection from severe SARS-CoV-2 infection is predictive of the observed protection in observational studies and RCTs (Figure 3A and B). A potential limitation in this analysis is that neutralisation titres were not directly measured in the epidemiological studies and thus predictions relied on an estimation of neutralisation titres based on vaccine type, variant, and time since vaccination. Therefore, we next sought to assess the utility of the published model of correlates of protection from severe COVID-19 using an approach that did not rely on estimating neutralising antibody titre. The published correlates model^4^ explicitly predicts a (non-linear) relationship between protection from symptomatic disease and protection from severe disease (red line in Figure 3C). That is, for any observed level of protection from symptomatic infection, the published model implicitly predicts a corresponding level of protection from severe COVID-19. This has the major advantage of being independent of any assumptions of the underlying neutralising antibody titres. Figure 3C shows the relationship between symptomatic and severe protection predicted from the correlates model (solid line and shaded 95% confidence intervals) and the data from a subset of 3 studies that reported protection against symptomatic and severe COVID-19 for comparable groups of subjects (Figure 1D). This subset of data included 178 observational measurements corresponding to 89 matched symptomatic and severe efficacy measurements. The data are shown on Figure 3C as points and associated 95% confidence intervals (whiskers). We observe that the real-world data points maintain the predicted relationship between symptomatic and severe protection (Spearman’s *ρ* = .63, p<.001), and note that 68/89 real world data points (90%) have a severe efficacy that lies within the 95% confidence intervals based on the corresponding symptomatic efficacy. We note that of the 21 reported points that lie outside of the model confidence intervals, 17 (81%) had predicted efficacies against severe disease above 97%. This is a range where it can be difficult to accurately measure efficacy in clinical studies. For all 17 of these datapoints the reported efficacy against severe disease was above 93%. These findings show that vaccine effectiveness against severe COVID-19 is not decoupled from effectiveness against symptomatic infection, but highly correlated and predictable based on the previously published relationship between neutralisation and protection^4^.

## Discussion

Neutralising antibodies are an important immune correlate of vaccine-mediated protection from symptomatic SARS-CoV-2 infection^4-7,14^. However, their role as a correlate of protection from severe SARS-CoV-2 infection has been less clear. The initial work identifying neutralising antibodies as a correlate of protection from symptomatic infection suggested lower titres of neutralising antibodies may be associated with protection from severe COVID-19. That is, while a level of neutralising antibodies equivalent to 20% of the GMT of early convalescent subjects (around 54 IU/ml) was associated with 50% protection from symptomatic infection, protection from severe infection was predicted to be achieved with a 6.5-fold lower titre (3.1% of convalescent, around 8 IU/ml)^4^. Unfortunately, the neutralisation level equivalent to 20% of convalescent titre is close to the detection limit in most assays reported in the phase 1 / 2 vaccine studies, while a titre equivalent to 3.1% of convalescent level is below the limit of detection in 5/7 of the reported assays^4^. This has led to a perception of ‘protection in the absence of neutralisation’^18^. However, this lack of detection arises largely because of the relatively high serum dilutions used in most *in vitro* assays, with a serum dilution of 1:10 or 1:20 being the lowest tested in most cases^19-21^. Thus, the relative preservation of protection from severe infection as well as the observed protection in the absence of detectable *in vitro* neutralisation, while perhaps not intuitive, were actually predicted by early studies of neutralising antibodies as correlates of protection^4^.

Although the association between neutralising antibodies and protection from symptomatic SARS-CoV-2 infection has been investigated in several settings^4-7,14,22^, protection from severe infection has heretofore remained more difficult to unravel. Here we aggregate the available epidemiological data to investigate whether the relationship between neutralising antibodies and protection from severe SARS-CoV-2 remains predictive across a diverse range of real-world scenarios of different vaccines, variants, and time since immunisation. Our analysis demonstrates that neutralising antibodies are indeed predictive of protection from severe COVID-19 across these different scenarios (Figure 3). Observations that vaccine protection against severe COVID-19 is relatively maintained against variants (compared to protection from symptomatic infection), combined with the fact that protection against symptomatic infection appears to wane faster than protection against severe COVID-19^8,9,23^ has led some to conclude that there is a ‘de-coupling’ in protection from symptomatic and severe disease^13^. However, this appears built on an expectation that protection will wane in parallel for different COVID-19 severity. Here we show that protection against severe COVID-19 is in fact strongly coupled with and predictable from the protection against symptomatic infection (albeit in a non-linear fashion) (Figure 3C), and that this relationship is consistent with predictions of the previously published model of the relationship between neutralising antibody titres and protection (Figure 3A and B)^4^.

That antibodies are capable of reducing the risk of severe COVID-19 is perhaps not surprising, since monoclonal antibody therapy administered in the first five days after symptom onset has been shown to reduce the risk of hospitalisation for severe SARS-CoV-2 by up to 85%, when administered in doses comparable to the neutralisation titres achieved in individuals vaccinated with an mRNA vaccine and/or booster (i.e. 7-fold the neutralisation titre found in the average convalescent individual)^24,25^. Indeed, some studies suggest that passive antibody administration may remain effective even later in infection when administered to seronegative subjects^26^. Together this suggests that spike-specific antibodies may play a mechanistic role in protection from progression from symptomatic to severe COVID-19.

This study has a number of limitations. Firstly, it aggregates the available epidemiological studies, which are heterogenous with respect to vaccines, variants, and study methodology (see Table S1). In addition, these real-world observational studies report vaccine effectiveness (in a non-randomised setting), whereas previous relationship between neutralisation titre and protection was derived from analysis of vaccine efficacy in randomised controlled trials^4^. We have previously shown that observational Test Negative Case Control studies (TNCC) tend to report a higher level of protection than seen in randomised trials^14^, although the effects of the different studies designs used in our analysis are unclear (Figure S1). Another limitation is that neutralising antibody titres were not directly measured in the study populations. Instead, neutralisation titres at different times and against different variants had to be estimated based on published parameters (see supplementary materials and Tables S3 and S4). However, in the absence of a widely accepted standard assay for *in vitro* neutralisation, it is unlikely that, even if neutralisation titres had been measured, they would have been comparable between studies^27^. Future studies directly measuring titres in exposed populations may be needed to directly validate our observations. However, given the low frequency of severe SARS-CoV-2 infection and the fact that the neutralisation levels associated with protection from severe COVID-19 are below detection in many assays, such studies may prove challenging.

The recent Omicron BA.1 and BA.2 waves across much of the world have illustrated the capacity of the virus to evade humoral immunity at the population level, with many previously effective vaccines showing low levels of protection from symptomatic Omicron variant infection^8^. Our analyses included data from 3 studies reporting on vaccine effectiveness against severe COVID-19 from the Omicron variant. We found that neutralising antibodies remain predictive of protection under this scenario^8,28,29^ (Figure S3, panels M and N, Spearman correlation *p*<.003 for both symptomatic and severe COVID-19). The relative maintenance of protection from hospitalisation and death have greatly reduced the public health burden of infection^29^. It will be increasingly important to understand disease severity as an endpoint in future epidemiological monitoring studies. In this context, identifying neutralising antibodies as a robust correlate of protection against severe COVID-19 is both timely and essential. In conclusion, we show that the relationship between neutralising antibody titres and protection holds across the spectrum of COVID-19 severity, and that neutralising antibody titres are predictive of protection against symptomatic as well as severe COVID-19.

## Methods

### Multiple Regression Fitting

To determine if vaccine effectiveness against severe COVID-19 was dependant on vaccine, variant and/or time since vaccination we fit a multiple linear regression model to vaccine effectiveness with vaccine and variant as categorical covariates, and time as a continuous covariate and interactions of these variables. The model with the lowest Akaike Information Criterion (AIC) was selected as the best model. The model with the lowest AIC was one that included vaccine type, variant and time since vaccination as well as an interaction between variant and time since vaccination, indicating variable wanning with time across variants. Therefore the resulting regression model was:

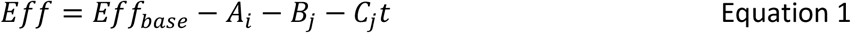

Where *A*_*i*_ is a vaccine specific adjustment for vaccine *i, B*_*j*_ is a variant specific adjustment for variant *j, C*_*j*_ is a variant specific parameter determining the change in effectiveness over time since vaccination (*t*). Values of these parameters are given in Table S2.

### Estimating mean neutralising antibody titres

We estimated the mean neutralising antibody titre that would be associated with each real-world effectiveness data point. This estimated neutralising antibody titre was based on:

1. The vaccine that was administered
2. The variant against which effectiveness is being measured
3. The time since vaccination
4. The dosing schedule for the vaccine
5. The timeframe over which efficacy was reported in the original phase 3 trials compared to the time frame measured in the extracted real-world data points.

We then combined these factors into an estimate for the mean neutralising antibody titres that would have been observed over the time period for that matches the reported effectiveness. Detailed equations describing how these factors were used to estimate neutralising antibodies are given in the Supplementary Materials.

### Determining confidence intervals using parametric bootstrapping

Confidence intervals of all estimates for neutralising antibody titres and predicted efficacies (shaded regions) in Figures 2, 3, S1-S4 were generated using parametric bootstrapping on the parameters with uncertainty in their estimation (as previously reported in reference^14^, detailed in supplementary methods using parameters in Tables S3 and S4).

### Statistical Analysis

All statistical comparisons were performed using R (version 4.0.2). Tests performed were Spearman’s rank correlations unless otherwise stated.

## Data Availability

Data and code are available upon request after publication.

## Ethics statement

This work was approved under the UNSW Sydney Human Research Ethics Committee (approval HC200242).

## Authorship Statement

DC, MPD and DSK contributed to study design. DC and MS performed the meta analysis and data extraction and curation. DC, AR, DSK and TES performed the data analysis. DC, MPD, DSK, SJK and SCS contributed to shaping the direction of the work. All authors contributed to the writing and reviewed and approved the final report.

## Data Availability Statement

Data and code are available upon request after publication.

## Acknowledgements

This work would not be possible without the many scientists who generously provided the published data analysed in this study, either through making the data directly available through the original publication or through providing it upon request. The authors thank these scientists for their contribution and the individual sources of data are indicated in the references and supplementary tables.

## Supplementary Figures

**Supplementary Figure 1:**
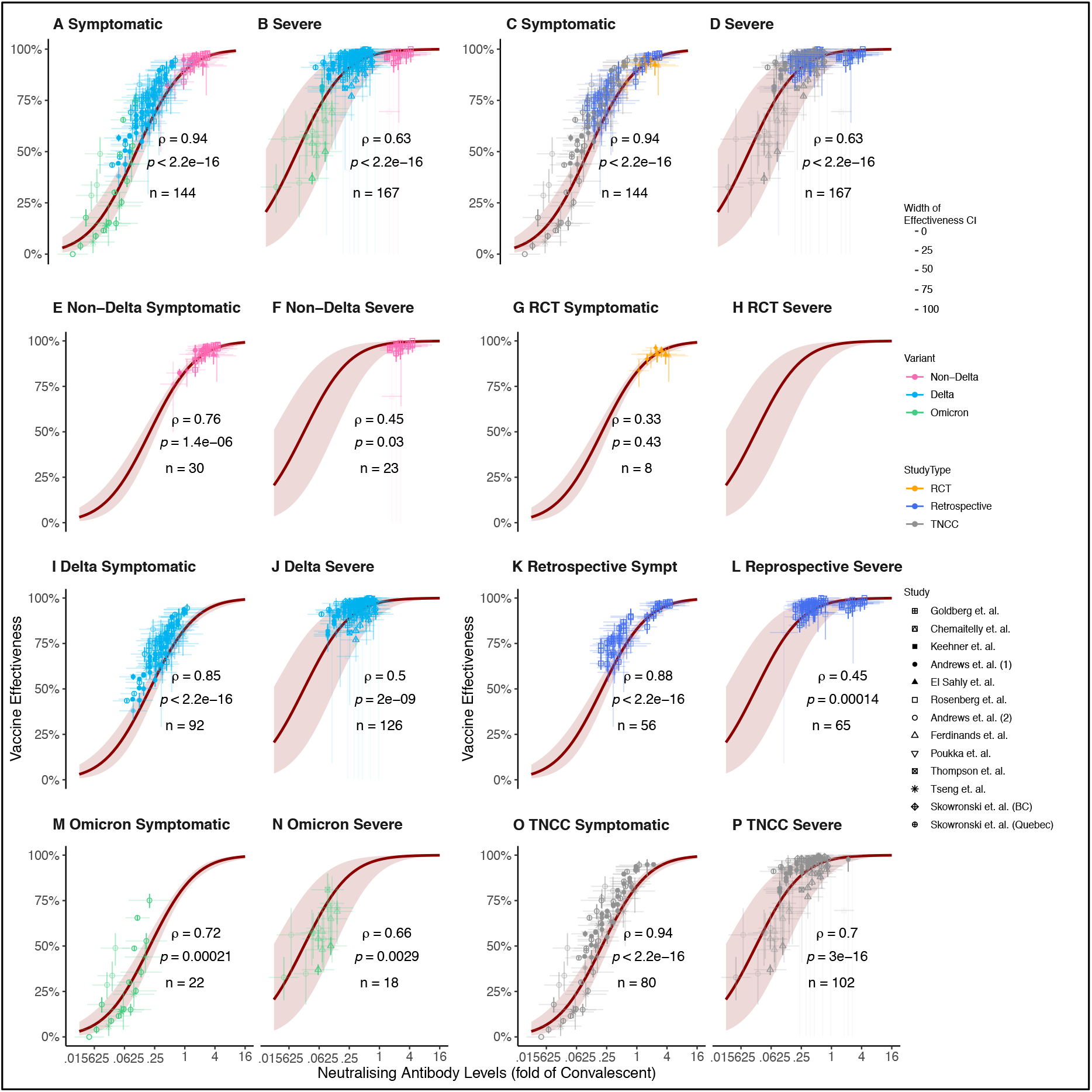
Correlation between estimated neutralising antibody titres (accounting for vaccine used, variant studied and time since vaccination) and clinical data for vaccine effectiveness against symptomatic and severe COVID-19, coloured by variant (left hand side, panels A,B,E,F,I,J,M,N) and study type (right hand side, panels C,D,G,H,K,L,O,P). Top row includes all data, bottom three rows include data split by variant (left two columns) and study type (right two columns). Numbers show ρ and p-value for Spearman correlations and the number of data-points shown in each plot. Note that our meta-analysis did not include any additional data for severe disease in a randomised control trial and so there is no extracted data shown in panel (H).

**Supplementary Figure 2:**
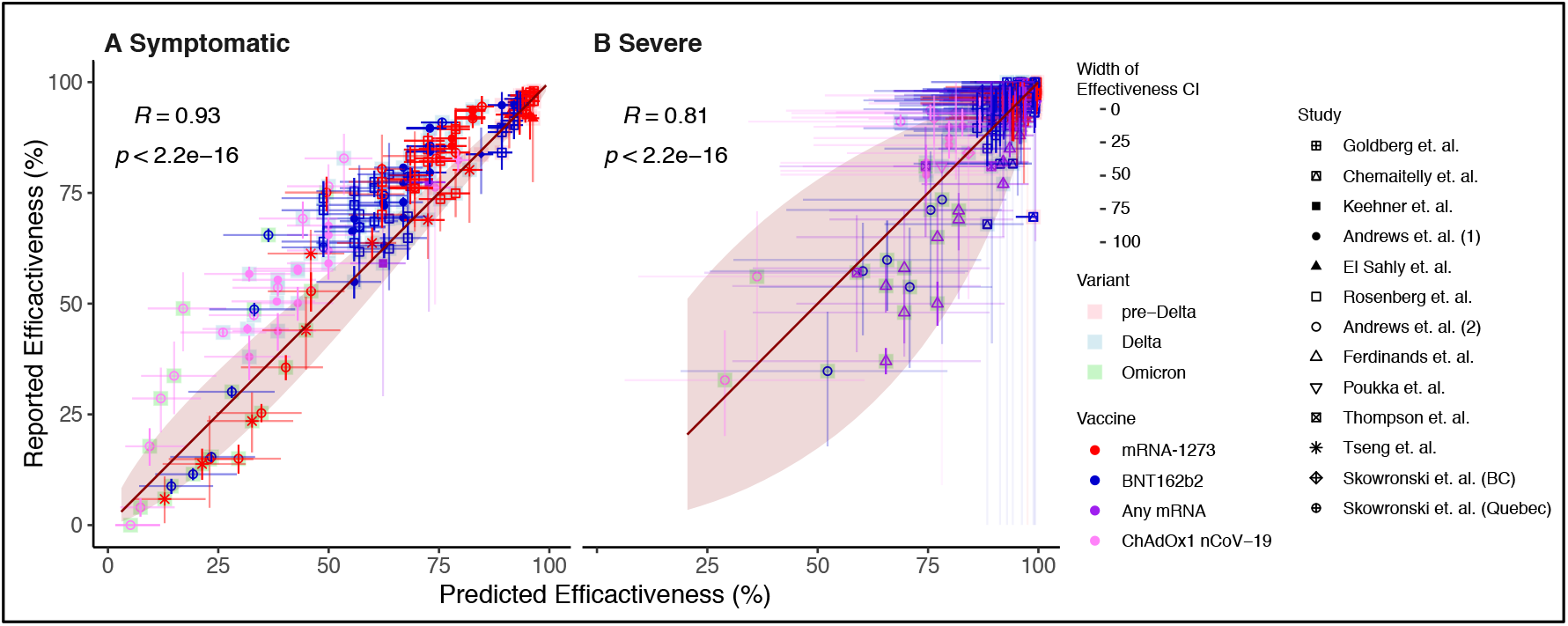
Comparison between predicted efficacy for a specific vaccine, variant and timepoint (x-axis) and the corresponding observed effectiveness estimate from the meta-analysis (y-axis) for (A) symptomatic and (B) severe COVID-19. Dark red line shows 1:1 relationship, and red band shows the 95% confidence intervals (as determined by parametric bootstrapping). Numbers show r-value for Pearson correlations.

**Supplementary Figure 3.**
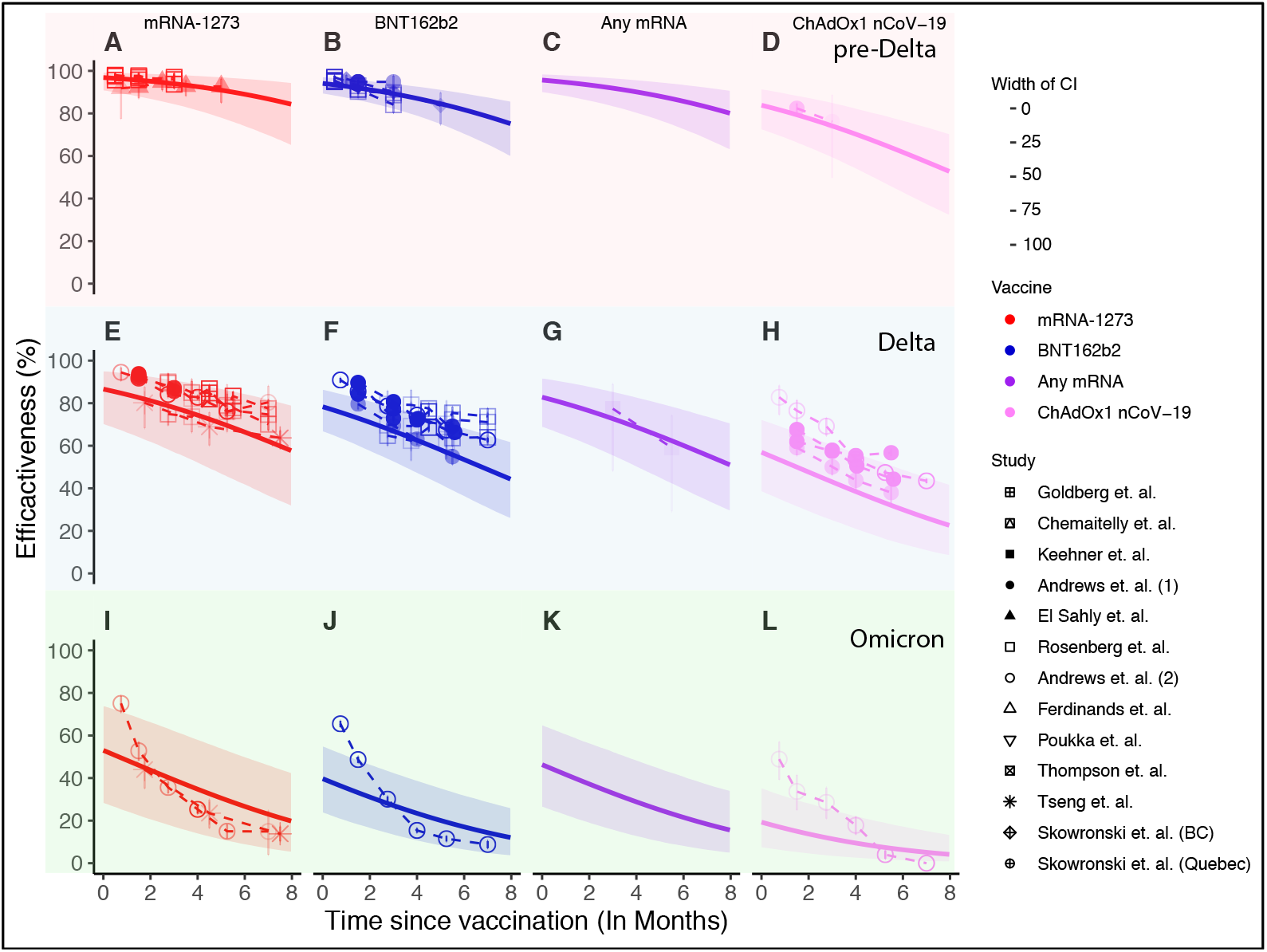
Model estimates (solid lines) and 95% confidence intervals (shaded area, as determined by parametric bootstrapping) for vaccine effectiveness against symptomatic SARS-CoV-2 infection with pre-Delta (top row, panels A-D), Delta (middle row, panels E-H) and Omicron (bottom row, panels I-L) variants. Data extracted from the clinical studies of vaccine effectiveness are overlaid as points (whiskers indicate 95% CI). Note that for panels C and K no effectiveness data was available. Model parameters and distributions are taken from references 4,14,17 and outlined in the supplementary methods.

**Supplementary Figure 4.**
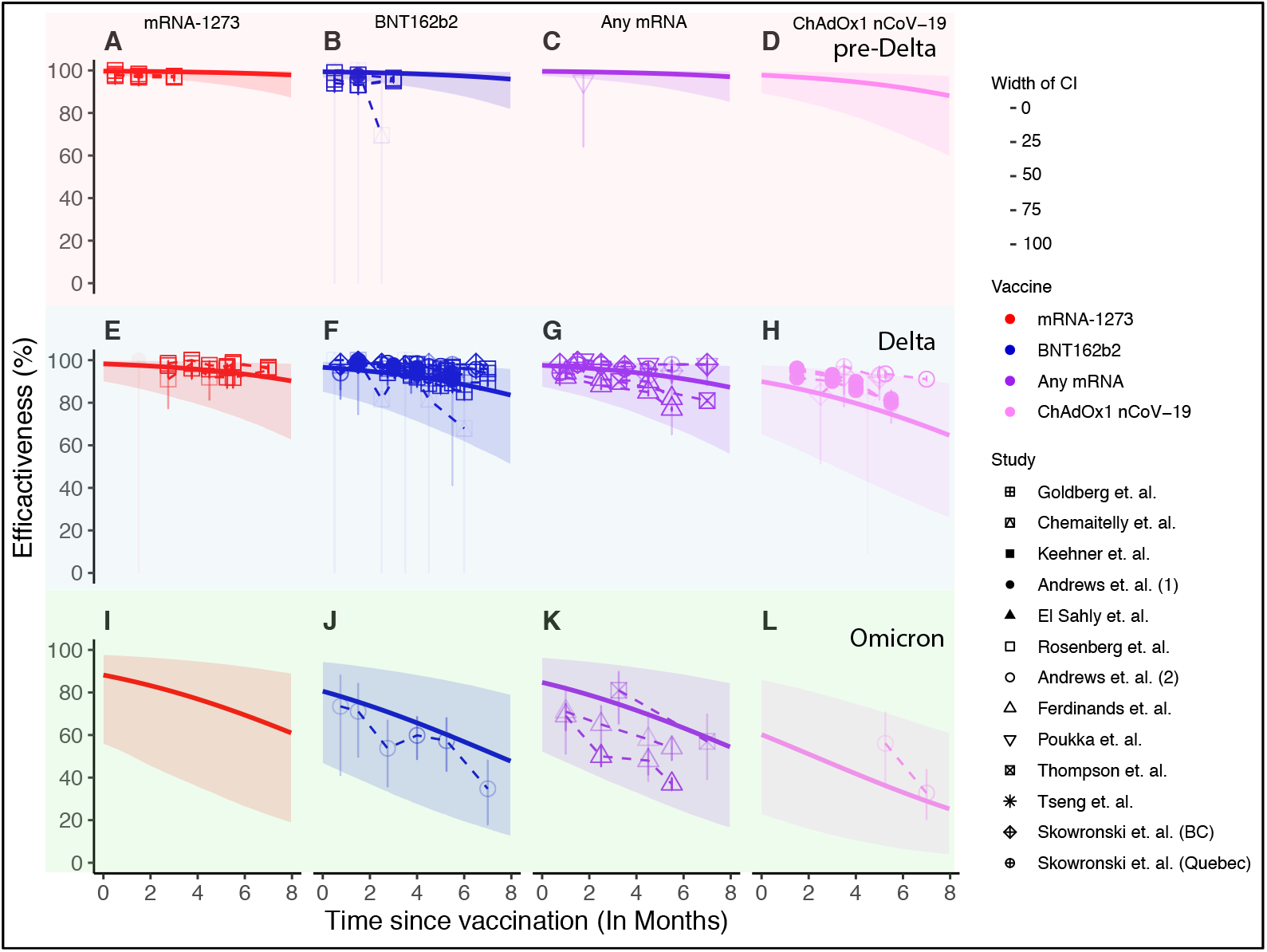
Model estimates (solid lines) and 95% confidence intervals (shaded area, as determined by parametric bootstrapping) for vaccine effectiveness against severe SARS-CoV-2 infection with pre-Delta (top row, panels A-D), Delta (middle row, panels E-H) and Omicron (bottom row, panels I-L) variants. Data extracted from the clinical studies of vaccine effectiveness are overlaid as points (whiskers indicate 95% CI). Note that for panels D and I no effectiveness data was available. Model parameters and distributions are taken from references 4,14,17 and outlined in the supplementary methods.

## References

1. Voysey M, Clemens SAC, Madhi SA, et al. Safety and efficacy of the ChAdOx1 nCoV-19 vaccine (AZD1222) against SARS-CoV-2: an interim analysis of four randomised controlled trials in Brazil, South Africa, and the UK. Lancet 2021; 397(10269): 99–111.

2. Polack FP, Thomas SJ, Kitchin N, et al. Safety and Efficacy of the BNT162b2 mRNA Covid-19 Vaccine. N Engl J Med 2020; 383(27): 2603–15.

3. Baden LR, El Sahly HM, Essink B, et al. Efficacy and Safety of the mRNA-1273 SARS-CoV-2 Vaccine. N Engl J Med 2021; 384(5): 403–16.

4. Khoury DS, Cromer D, Reynaldi A, et al. Neutralizing antibody levels are highly predictive of immune protection from symptomatic SARS-CoV-2 infection. Nat Med 2021; 27(7): 1205–11.

5. Feng S, Phillips DJ, White T, et al. Correlates of protection against symptomatic and asymptomatic SARS-CoV-2 infection. Nat Med 2021; 27(11): 2032–40.

6. Gilbert PB, Montefiori DC, McDermott AB, et al. Immune correlates analysis of the mRNA-1273 COVID-19 vaccine efficacy clinical trial. Science 2022; 375(6576): 43–50.

7. Bergwerk M, Gonen T, Lustig Y, et al. Covid-19 Breakthrough Infections in Vaccinated Health Care Workers. N Engl J Med 2021; 385(16): 1474–84.

8. Andrews N, Stowe J, Kirsebom F, et al. Covid-19 Vaccine Effectiveness against the Omicron (B.1.1.529) Variant. N Engl J Med 2022; 386(16): 1532–46.

9. Andrews N, Tessier E, Stowe J, et al. Duration of Protection against Mild and Severe Disease by Covid-19 Vaccines. N Engl J Med 2022; 386(4): 340–50.

10. Chemaitelly H, Tang P, Hasan MR, et al. Waning of BNT162b2 Vaccine Protection against SARS-CoV-2 Infection in Qatar. N Engl J Med 2021; 385(24): e83.

11. Tartof SY, Slezak JM, Puzniak L, et al. Durability of BNT162b2 vaccine against hospital and emergency department admissions due to the omicron and delta variants in a large health system in the USA: a test-negative case-control study. Lancet Respir Med 2022.

12. Tseng HF, Ackerson BK, Luo Y, et al. Effectiveness of mRNA-1273 against SARS-CoV-2 Omicron and Delta variants. Nat Med 2022.

13. Madhi SA, Ihekweazu C, Rees H, Pollard AJ. Decoupling of omicron variant infections and severe COVID-19. Lancet 2022; 399(10329): 1047–8.

14. Cromer D, Steain M, Reynaldi A, et al. Neutralising antibody titres as predictors of protection against SARS-CoV-2 variants and the impact of boosting: a meta-analysis. Lancet Microbe 2022; 3(1): e52–e61.

15. Khoury DS, Steain M, Triccas JA, Sigal A, Davenport MP, Cromer D. A meta-analysis of Early Results to predict Vaccine efficacy against Omicron. medrxiv 2021: doi:10.1101/2021.12.13.21267748.

16. Goel RR, Painter MM, Apostolidis SA, et al. mRNA vaccines induce durable immune memory to SARS-CoV-2 and variants of concern. Science 2021; 374(6572): abm0829.

17. Cele S, Jackson L, Khoury DS, et al. Omicron extensively but incompletely escapes Pfizer BNT162b2 neutralization. Nature 2022; 602(7898): 654–6.

18. Kalimuddin S, Tham CYL, Qui M, et al. Early T cell and binding antibody responses are associated with COVID-19 RNA vaccine efficacy onset. Med (N Y) 2021; 2(6): 682–8 e4.

19. Jackson LA, Anderson EJ, Rouphael NG, et al. An mRNA Vaccine against SARS-CoV-2 -Preliminary Report. N Engl J Med 2020; 383(20): 1920–31.

20. Keech C, Albert G, Cho I, et al. Phase 1-2 Trial of a SARS-CoV-2 Recombinant Spike Protein Nanoparticle Vaccine. N Engl J Med 2020; 383(24): 2320–32.

21. Walsh EE, Frenck RW, Jr., Falsey AR, et al. Safety and Immunogenicity of Two RNA-Based Covid-19 Vaccine Candidates. N Engl J Med 2020; 383(25): 2439–50.

22. Earle KA, Ambrosino DM, Fiore-Gartland A, et al. Evidence for antibody as a protective correlate for COVID-19 vaccines. Vaccine 2021; 39(32): 4423–8.

23. Rosenberg ES, Dorabawila V, Easton D, et al. Covid-19 Vaccine Effectiveness in New York State. N Engl J Med 2022; 386(2): 116–27.

24. Stadler E, Chai KL, Schlub TE, et al. Determinants of passive antibody effectiveness in SARS-CoV-2 infection. medrxiv 2022: doi:10.1101/2022.03.21.22272672.

25. Gupta A, Gonzalez-Rojas Y, Juarez E, et al. Early Treatment for Covid-19 with SARS-CoV-2 Neutralizing Antibody Sotrovimab. N Engl J Med 2021; 385(21): 1941–50.

26. Group RC. Casirivimab and imdevimab in patients admitted to hospital with COVID-19 (RECOVERY): a randomised, controlled, open-label, platform trial. Lancet 2022; 399(10325): 665–76.

27. Khoury DS, Schlub TE, Cromer D, et al. Correlates of protection, thresholds of protection, and immunobridging in SARS-CoV-2 infection. medrxiv 2022: doi:10.1101/2022.06.05.22275943.

28. Thompson MG, Natarajan K, Irving SA, et al. Effectiveness of a Third Dose of mRNA Vaccines Against COVID-19-Associated Emergency Department and Urgent Care Encounters and Hospitalizations Among Adults During Periods of Delta and Omicron Variant Predominance -VISION Network, 10 States, August 2021-January 2022. MMWR Morb Mortal Wkly Rep 2022; 71(4): 139–45.

29. Ferdinands JM, Rao S, Dixon BE, et al. Waning 2-Dose and 3-Dose Effectiveness of mRNA Vaccines Against COVID-19-Associated Emergency Department and Urgent Care Encounters and Hospitalizations Among Adults During Periods of Delta and Omicron Variant Predominance -VISION Network, 10 States, August 2021-January 2022. MMWR Morb Mortal Wkly Rep 2022; 71(7): 255–63.

