## Supplementary Table 1 for "Neutralising antibodies predict protection from severe COVID-19"

| Study | Vaccine | Timeframe postvaccination | Measure/s of effectiveness | Age groups included | Variants analysed | Study Type | Country | Data derived from |
|---|---|---|---|---|---|---|---|---|
| Tartof et. al.[†1] | BNT162b2[A] | 0.5 – 7.25 | i) PCR positive<br>ii) Hospitalisation[B] | 12-15<br>16-44<br>45-64<br>≥65 | pre-Delta<br>Delta | Retrospective cohort study | USA | Appendix tables 6-8 |
| Goldberg et. al.[†2] | BNT162b2[A] | 1.5-6.5 months | i) PCR positive[C]<br>ii) Severe disease[D] | 16-39<br>40-59<br>≥60 | Delta | Retrospective cohort study | Israel | Table S7 |
| Chemaitelly et. al.[†3] | BNT162b2 | 0.5-≥7 months | i) PCR positive<br>ii) Hospitallisation (Severe disease Critical Disease And Fatal) | any | Alpha, Beta[E], Delta | Test negative case control | Qatar | Table 2 |
| Keehner et. al.[4] | BNT162b2 or mRNA-1273 | 1-7 months | PCR positive and > 1 symptom | ≥18 | Delta[F] | Retrospective cohort study | USA | Calculated based on attack rates for July given in text |
| Andrews et. al.[5] | BNT162b2<br>ChAdOx1 nCoV-19<br>mRNA-1273 | 0.5-≥9 months | i) PCR confirmed symptomatic disease<br>ii) Hospitalisation<br>iii) Death | ≥16<br>≥65<br>40-64<br>16-39 | Alpha<br>Delta | Test negative case control | England | Table 1<br>Table 2<br>Table S10<br>Table S11<br>Table S12 |
| El Sahly et. al.[6] | mRNA-1273 | 0.5-≥8 months | i) Prevention of illness<br>ii) Prevention of severe disease | ≥18-<65<br>≥65 | Ancestral | Randomised controlled trial | USA | Supplementary Table S30 |

| | | | | | | | | |
|---|---|---|---|---|---|---|---|---|
| Thomas et. al.[7] | BNT162b2 | 0.25-≥4 months | i) Laboratory confirmed disease (≥ 1 symptom) | 12-15<br>≥16 | pre-Delta | Randomised controlled trial | USA<br>Argentina<br>Brazil<br>South Africa<br>Germany<br>Turkey | Figure 2 |
| Rosenberg et. al.[8] | BNT162b2<br>mRNA-1273 | 0-8 months | i) Laboratory confirmed disease<br>ii) Hospitalisation | 18-49<br>50-64<br>≥65 | pre-Delta<br>Delta | Retrospective cohort study | USA | Tables 2 and 3 |
| Andrews et. al.[9,10] | BNT162b2<br>ChAdOx1 nCoV-19<br>mRNA-1273 | 0.5-~25 months | i) PCR confirmed symptomatic disease<br>ii) Hospitalisation | ≥18 | Delta<br>Omicron | Test negative case control | England | Table 3 |
| Ferdinands et. al.[11] | mRNA vaccines | 0.5-25 months | i) PCR confirmed hospital presentation<br>ii) PCR confirmed hospital admission | ≥18 | Delta<br>Omicron | Test negative case control | USA | Table 2 |
| Bruxvoort et. al.[†12] | mRNA-1273 | 0.5-6 months | PCR confirmed infection | ≥18 | Delta | Test negative case control | USA | Extracted from Figure 2 |
| Poukka et. al.[†13] | mRNA vaccines<br>ChAdOx1 nCoV-19 | 0-8 months | i) Laboratory confirmed infection<br>ii) Hospitalisation | 16-69 | pre-Delta<br>Delta | Retrospective cohort study | Finland | Supplementary table 2 and 3 |
| Tseng et. al.[14] | mRNA-1273 | 0.5-25 months | i) Infection ii) Hospitalisation | ≥18 | Delta<br>Omicron | Test negative case control | USA | Table 2 |
| Skowronski et. al.[†15] | BNT162b2 | 0.5-11.5 months | i) PCR positive infection | ≥18 | Delta | Test negative case control | Canada | Supplementary table 14 and 15 |

| | | | | | | | | |
|---|---|---|---|---|---|---|---|---|
| | ChAdOx1-nCoV-19<br>mRNA-1273 | | ii) hospitalisation | | | | | |
| Thompson et. al.[16] | mRNA vaccines | 0.5-25 months | Hospitalisation | ≥18 | Delta<br>Omicron | Test negative case control | USA | Table 2 |

*Supplementary Table 1 Data sources for Efficacy Data*

[A] Fully vaccinated = 7 days + post second dose
[B] Hospitalisation data was not split by variant
[C] PCR positive efficacy included for ages >16 years
[D] Severe disease efficacy included for ages >40 years
[E] Effectiveness against Beta infections were not included in this analysis
[F] Infections were >95% Delta in timeframe analysed
[†] Denotes studies that included efficacy against confirmed infection without reporting on symptoms. These data were not included in the final analysis.

**References:**


1. Tartof SY, Slezak JM, Fischer H, et al. Effectiveness of mRNA BNT162b2 COVID-19 vaccine up to 6 months in a large integrated health system in the USA: a retrospective cohort study. *Lancet* 2021; **398**(10309): 1407-16.
2. Goldberg Y, Mandel M, Bar-On YM, et al. Waning Immunity after the BNT162b2 Vaccine in Israel. *N Engl J Med* 2021; **385**(24): e85.
3. Chemaitelly H, Tang P, Hasan MR, et al. Waning of BNT162b2 Vaccine Protection against SARS-CoV-2 Infection in Qatar. *N Engl J Med* 2021; **385**(24): e83.
4. Keehner J, Horton LE, Binkin NJ, et al. Resurgence of SARS-CoV-2 Infection in a Highly Vaccinated Health System Workforce. *N Engl J Med* 2021; **385**(14): 1330-2.
5. Andrews N, Tessier E, Stowe J, et al. Duration of Protection against Mild and Severe Disease by Covid-19 Vaccines. *N Engl J Med* 2022; **386**(4): 340-50.
6. El Sahly HM, Baden LR, Essink B, et al. Efficacy of the mRNA-1273 SARS-CoV-2 Vaccine at Completion of Blinded Phase. *N Engl J Med* 2021; **385**(19): 1774-85.
7. Thomas SJ, Moreira ED, Jr., Kitchin N, et al. Safety and Efficacy of the BNT162b2 mRNA Covid-19 Vaccine through 6 Months. *N Engl J Med* 2021; **385**(19): 1761-73.
8. Rosenberg ES, Dorabawila V, Easton D, et al. Covid-19 Vaccine Effectiveness in New York State. *N Engl J Med* 2022; **386**(2): 116-27.
9. Andrews N, Stowe J, Kirsebom F, et al. Covid-19 Vaccine Effectiveness against the Omicron (B.1.1.529) Variant. *N Engl J Med* 2022; **386**(16): 1532-46.
10. Andrews N, Stowe J, Kirsebom F, et al. Effectiveness of COVID-19 vaccines against the Omicron (B.1.1.529) variant of concern. *medrxiv* 2021: doi:10.1101/2021.12.14.21267615.
11. Ferdinands JM, Rao S, Dixon BE, et al. Waning 2-Dose and 3-Dose Effectiveness of mRNA Vaccines Against COVID-19-Associated Emergency Department and Urgent Care Encounters and Hospitalizations Among Adults During Periods of Delta and Omicron Variant Predominance - VISION Network, 10 States, August 2021-January 2022. *MMWR Morb Mortal Wkly Rep* 2022; **71**(7): 255-63.
12. Bruxvoort KJ, Sy LS, Qian L, et al. Effectiveness of mRNA-1273 against delta, mu, and other emerging variants of SARS-CoV-2: test negative case-control study. *BMJ* 2021; **375**: e068848.
13. Poukka E, Baum U, Palmu AA, et al. Cohort study of Covid-19 vaccine effectiveness among healthcare workers in Finland, December 2020 - October 2021. *Vaccine* 2022; **40**(5): 701-5.
14. Tseng HF, Ackerson BK, Luo Y, et al. Effectiveness of mRNA-1273 against SARS-CoV-2 Omicron and Delta variants. *Nat Med* 2022; **28**(5): 1063-71.
15. Skowronski DM, Febriani Y, Ouakki M, et al. Two-dose SARS-CoV-2 vaccine effectiveness with mixed schedules and extended dosing intervals: test-negative design studies from British Columbia and Quebec, Canada. *medrxiv* 2021.
16. Thompson MG, Natarajan K, Irving SA, et al. Effectiveness of a Third Dose of mRNA Vaccines Against COVID-19-Associated Emergency Department and Urgent Care Encounters and Hospitalizations Among Adults During Periods of Delta and Omicron Variant Predominance - VISION Network, 10 States, August 2021-January 2022. *MMWR Morb Mortal Wkly Rep* 2022; **71**(4): 139-45.
