## Supplementary Methods for "Neutralising antibodies predict protection from severe COVID-19"

*Data on vaccine effectiveness*

We searched the available literature to identify studies that reported data on vaccine effectiveness against (i) a defined clinical endpoint (ii) for an identifiable variant, (ii) for a single vaccine (or vaccine type), (iii) over an identified time since vaccination, and (iv) for which data was either provided in or readily extractable from the original publication (see Table S1). We identified 15 published studies that met these criteria, which collectively provided over 311 individual data points on vaccine effectiveness. We focused on efficacy for three of the main vaccines used in primary vaccination regimes – mRNA-1273, BNT162b2 and ChAdOx1-nCoV-19, and on efficacy against symptomatic and severe COVID-19 disease outcomes. These studies are detailed in Table S1.

*Extraction of vaccine effectiveness data*

Data was extracted from identified papers either directly from tables included within the publication, or else was extracted from figures using WebPlotDigitizer [1]. The data used for each study is indicated in Table S1.

*Multiple Regression Fitting*

As described in the main manuscript, we used a multiple linear regression model on vaccine effectiveness with vaccine, variant and time since vaccination as independent variables.

*Estimating Neutralising Antibody Titres*

To estimate the mean neutralising antibody titre that would be associated with a given real-world effectiveness estimate, we account for a number of influencing factors. These are:

1) The vaccine that was administered
2) The variant against which effectiveness is being measured
3) The time since vaccination
4) The dosing schedule for the vaccine
5) The timeframe over which efficacy was reported in the original phase 3 trials compared to the time frame measured in the extracted real-world data points.

These factors were included in the estimate as described below.

*1) Accounting for the vaccine administered*

We have previously estimated the $\log_{10}$ of the starting neutralisation titres observed for a number of vaccines (relative to the geometric mean convalescent titres) using data from phase 1/2 trials for 7 different vaccines [2]. We use these estimates to estimate the peak mean neutralising antibody titres that would have been observed for each vaccine against the wild type variant, using the same dosing schedule as in the relevant phase I/II trials. These estimates are denoted by $\mu_i$ (for vaccine $i$) and are given in Table S3.

*2) Accounting for the variant*

We have previously estimated the drop in neutralisation titres observed for a number of VoC by combining data from 17 different studies across 5 different vaccines and 5 different variants. This work showed that the fold drop in neutralisation titre (for a given variant) is independent of the vaccine administered [3]. In addition, we have also studied the drop in neutralisation titre to the Omicron variant [4]. Here we use the fold drop estimated from our previous meta analysis for the Delta variant (3.9-fold) [3], and the fold drop estimated by Cele et. al. for the Omicron variant (22-fold) [4]. We assume there is no change to the neutralisation titres compared to the ancestral variant for the pre-Delta variants. The fold drops used are denoted by $f_j$ (for variant $j$) and are specified in Table S3.

*3) Accounting for the time since vaccination*

To account for waning neutralising antibody levels over the period since vaccination, we assumed that neutralising antibody levels decay exponentially over the trial period according to the formula

$$N_{ab}(t) = N_{ab}(0)e^{-\delta t}. \qquad \text{Equation S1}$$

Where $\delta$ corresponds to a half-life of 108 days (estimated in[2], using data from [5]). The distribution for this rate is specified in Table S2.

*4) Adjusting for the vaccine dosing schedule*

The phase 2 neutralising antibody data used to parameterise the original correlates model for ChAdOx-nCoV-1 was based on a 3 week interval between the first and second vaccine dose. This dosing schedule was subsequently increased to 6-12 weeks in the majority of real world scenarios, resulting in a 50%-90% increase in neutralising antibody levels[6]. In order to account for this discrepancy between the real world implementation and the clinical trial antibody levels we adjusted the neutralising antibody levels for ChAdOx1-nCov vaccinees upwards by a factor of 1.59 fold, to match the estimated increase seen with a 9 week dosing schedule[6]. Therefore we adjusted the antibody levels further by a factor of $\phi_i$ where

$$\phi_i = \begin{cases} 1.59 & \text{for ChAdOx-nCoV-1} \\ 0 & otherwise \end{cases} \qquad \text{Equation S2}$$

*5) Adjusting for duration of follow-up and duration of the original phase 3 trials.*

The correlates model[2] was originally fitted to the peak antibody titres seen in the Phase I/II vaccine trials (approximately 2-3 weeks after administration of the final vaccine dose) and the reported vaccine efficacy over the duration of the phase 3 trial randomised control trial (median length of follow up in most phase of the 3 trials was 2 months in line with FDA requirements[7]). Thus, it essentially reports the relationship between peak neutralising antibody titres at the start of follow up and vaccine protection over the subsequent (approximately) two months of follow up. However, the vaccine effectiveness studies analysed here reported protection over differing periods ranging from 2 weeks to 4 months (i.e. not 2 months in most cases). Differences in the duration of follow-up may confound results due to wanning protection over the course of longer studies. This effect was accounted for by estimating the mean neutralisation level and associated predicted efficacy at the mid-point of follow-up time periods, and using this mid-point in Figures 2, S3 and S4. This mid-point estimation uses the expected antibody decay kinetics from equation S1, to adjust the prediction of efficacy from Khoury et. al.[2], which are based on neutralisation titres at the start of the follow up period. This is essentially an adjustment by a factor of $e^{\delta T}/_2$ (where $\delta$ is the neutralising antibody decay rate, and $T$ is the trial length). Figures 3A and 3B similarly plot the reported vaccine efficacy at the mid-point of the reported time interval against the estimated neutralising antibody titre (adjusted as above) that should be used to predict efficacy at the mid-point of the interval.

Where a time interval is reported as “more than X weeks / months” we calculate the instantaneous adjusted neutralising antibody titres and associated vaccine effectiveness one month after the lower time bound on this group. i.e. if a study reported efficacy at “more than 5 months” we would correlate this with estimated values at 6 months – one month more than the lower bound of 5 months.

*Combined estimate of neutralising antibody levels*

The overall neutralising antibody titre is then calculated by incorporating each of the factors described above. Therefore, the neutralising antibody levels calculated for vaccine $i$ against variant $j$ at time $t$ after vaccination, $N_{ab}(i,j,t)$ is given by:

$$N_{ab}(i,j,t) = 10^{\left(\mu_i - f_j + \phi_i + \log_{10} e^{-\delta\left(t - \frac{T}{2}\right)}\right)}. \qquad \text{Equation S3}$$

Where $\mu_i$ represents the mean of the $\log_{10}$ of the neutralising antibody levels against ancestral virus for subjects vaccinated with vaccine $i$, $f_j$ represents the fold drop in neutralising antibody titre for variant $j$ (compared to ancestral virus), $\delta$ is the decay rate of neutralising antibodies, and $T$ is the average length of the original phase III trials.

As all of the factors determining the neutralisation titre for a given vaccine regimen at a given time each contain their own confidence bands, the cumulative effect of these confidence bands is used to calculate the overall uncertainty in the neutralising antibody levels (see next section and Table S3).

*Predicting vaccine effectiveness using the previously published correlates model*

We have previously developed and fitted a model correlating neutralising antibody titres (immunogenicity data taken from phase I/II trials) to vaccine efficacy (protective efficacy taken from phase III trials) against symptomatic and severe SARS-CoV-2 infection[2]. This published model was parameterised using data from seven published studies of vaccine efficacy along with data on protection from COVID-19 after previous infection. Specifically, vaccine effectiveness, VE, is defined as:

$$VE(\mu_i, f_j) = \int_{-\infty}^{\infty} N(x, \mu_i - f_j, \sigma) \frac{1}{1+e^{-k(x-x_{50})}} \, dx \qquad \text{Equation S4}$$

Where $\sigma = 0.46$, $k = 3.1$ and $x_{50} = \log_{10} 0.2$ for symptomatic infection and $x_{50} = \log_{10} 0.03$ for severe infection[2,8] (Table S4). Where, as above, $\mu_i$, represents the mean of the $\log_{10}$ of the neutralising antibody titres for vaccine $i$ against the ancestral strain of the virus, and $f_j$ represents the fold decrease in neutralising antibody titres for variant *j*. Values of these parameters were collected from the literature as outlined in Tables S3 and S4. In this work, references to the “correlates model” refer to the use of this model, as originally published and parameterised. I.e. the parameters of this model were not re-estimated or fitted in this study, but were used as originally reported[2].

*Determining confidence intervals using parametric bootstrapping*

Confidence intervals of all estimates for neutralising antibody titres and predicted efficacies (shaded regions) in Figures 2, 3, S1-S4 were generated using parametric bootstrapping on the parameters with uncertainty in their estimation (as previously reported in reference[3], parameters given in Tables S3 and S4) as follows. For any time point along the x-axis in Figures 2, S3 and S4, or for the horizontal placement of data points in Figures 3A and B, the mean neutralising antibody titre was first estimated using equation S3. Then the confidence bands of the neutralising antibody titres was estimated by repeatedly using equation S3 to re-estimate the neutralising antibody titre, while sampling parameters from the distributions given in Table S3.

Subsequently, for any neutralisation ratio either calculated above, or for a position on the x-axis in Figure 3A or B, equation S4 was first used to estimate the mean corresponding protective efficacy. Then the distribution of efficacies was estimated by repeating the efficacy calculation with equation S4, using parameter values drawn randomly from distributions according to their standard error or covariance matrix (normal and bivariant normal distributions respectively in Table S4).

Sampling was performed 100,000 times and the lower confidence bound was estimated from at the 2.5% percentile, while the upper confidence bound was taken from the 97.5% percentile.

| **Parameter** | **Description** | **Vaccine / Variant** | **Effectiveness reduction value (95% CI)*** |
|---|---|---|---|
| $A_i$ | Vaccine Specific Efficacy difference | mRNA-1273 | reference |
| | | Any mRNA | 2.3 (-1.7-6.4) |
| | | BNT162b2 | 4.5 (1.1-7.8) |
| | | ChAdOx1 nCov-19 | 8.7 (4.4-13) |
| $B_j$ | Variant Specific Efficacy difference | pre-Delta | reference |
| | | Delta | 4.7 (–2.2-11.7) |
| | | Omicron | 27.2 (18.8-35.5) |
| $C_j$ | Loss in efficacy per month since vaccination | pre-Delta | 6.0 (2.5-9.5) |
| | | Delta | 1.2 (0.5-2.0) |
| | | Omicron | 4.4 (3.1-5.6) |

*Table S2 Parameters estimated in the multiple regression model fitting for severe COVID-19. *Note that a positive value indicates a lower estimated efficacy as the coefficients of equation 1 have a negative sign in front of them.*

| Parameter | Description | Vaccine / Variant | Mean Value | Distribution | Reference |
|---|---|---|---|---|---|
| $\mu_i$ | Starting neutralising antibody levels (as a fold of convalescent) | mRNA-1273 | $\log_{10} 4.1$ | $N(\log_{10} 4.1\,, .006)$ | [2] |
| | | BNT162b2 | $\log_{10} 2.4$ | $N\,(\log_{10} 2.4\,, .01)$ | [2] |
| | | ChAdOx1 nCov-19 | $\log_{10} 0.8$ | $N(\log_{10} 0.8\,, .018)$ | [2] |
| | | Any mRNA | $\log_{10} 3.1$ | $N(\log_{10} 3.1\,, .023)$ | (geometric mean of mRNA-1273 and BNT162b2) |
| $f_j$ | Fold change in neutralisation titre against variant | Delta | $-\log_{10} 3.9$ | $N(-\log_{10} 3.9\,, .003)$ | [3] |
| | | Omicron | $-\log_{10} 22$ | $N(-\log_{10} 22\,, .005)$ | [4] |
| $\delta$ | Neutralising antibody decay rate | N/A | $\ln 2/108$ | $N(6.42, .001) * 10^{-3}$ | [2,5] |
| $T$ | Phase 3 clinical trial length | N/A | 60 days | N/A | [9-11] |
| $\phi_i$ | Dosage adjustment for different ChAdOx1-nCoV-19 dose interval | ChAdOx-nCoV-1 | $\log_{10} 1.59$ | N/A | [6] |
| | | Other vaccines | 0 | N/A | N/A |

*Table S3 Parameters used in estimating neutralisation titre. The GMT value used for mRNA is the geometric mean value of mRNA-1273 and BNT162b2 values. Note that all these parameters were estimated from previously published work.*

| Parameter | Description | Endpoint | Mean Value | Distribution | Reference |
|---|---|---|---|---|---|
| $\sigma$ | | N/A | 0.46 | $N(0.465{,}0.022)$ | [2] |
| $k$ | Hill coefficient* | Symptomatic | $e^{1.13}$ | $e^{N(1.13,.031)}$ | [2] |
| | | Severe | $e^{1.12}$ | $e^{N(1.12,.03)}$ | [2] |
| $x_{50}$ | IC50 for protection against disease* | Symptomatic | $\log_{10} 0.20$ | $N(\log_{10} 0.20\,, 0.006)$ | [2] |
| | | Severe | $\log_{10} 0.03$ | $N(\log_{10} 0.03\,, 0.099)$ | [2] |

*Table S4 Model parameters used in estimating the relationship between neutralising antibody titre and protection from COVID-19 (taken from Khoury et. al. [2]) *Note that the hill coefficient and IC50 parameters are selected from a bivariate normal distribution with covariance matrix given by* $C = \begin{pmatrix} .031 & .011 \\ .011 & .006 \end{pmatrix}$ *for symptomatic protection and* $C = \begin{pmatrix} .03 & .03 \\ .03 & .099 \end{pmatrix}$ *for severe protection.*

### References


1. Rohatgi A. WebPlotDigitizer. 4.5 ed. Pacifica, California, USA.
2. Khoury DS, Cromer D, Reynaldi A, et al. Neutralizing antibody levels are highly predictive of immune protection from symptomatic SARS-CoV-2 infection. *Nat Med* 2021; **27**(7): 1205-11.
3. Cromer D, Steain M, Reynaldi A, et al. Neutralising antibody titres as predictors of protection against SARS-CoV-2 variants and the impact of boosting: a meta-analysis. *Lancet Microbe* 2022; **3**(1): e52-e61.
4. Cele S, Jackson L, Khoury DS, et al. Omicron extensively but incompletely escapes Pfizer BNT162b2 neutralization. *Nature* 2022; **602**(7898): 654-6.
5. Dan JM, Mateus J, Kato Y, et al. Immunological memory to SARS-CoV-2 assessed for up to 8 months after infection. *Science* 2021; **371**(6529).
6. Voysey M, Costa Clemens SA, Madhi SA, et al. Single-dose administration and the influence of the timing of the booster dose on immunogenicity and efficacy of ChAdOx1 nCoV-19 (AZD1222) vaccine: a pooled analysis of four randomised trials. *Lancet* 2021; **397**(10277): 881-91.
7. FDA. Emergency Use Authorization for Vaccines to Prevent COVID-19. 31 March 2022. https://www.fda.gov/media/142749/download (accessed 8 June 2022 2022).
8. Khoury DS, Cromer D, Reynaldi A, et al. R Code: Neutralizing antibody levels are highly predictive of immune protection from symptomatic SARS-CoV-2 infection. 2021/05/19 ed. https://github.com/InfectionAnalytics/COVID19-ProtectiveThreshold: Github; 2021.
9. Voysey M, Clemens SAC, Madhi SA, et al. Safety and efficacy of the ChAdOx1 nCoV-19 vaccine (AZD1222) against SARS-CoV-2: an interim analysis of four randomised controlled trials in Brazil, South Africa, and the UK. *Lancet* 2021; **397**(10269): 99-111.
10. Polack FP, Thomas SJ, Kitchin N, et al. Safety and Efficacy of the BNT162b2 mRNA Covid-19 Vaccine. *N Engl J Med* 2020; **383**(27): 2603-15.
11. Baden LR, El Sahly HM, Essink B, et al. Efficacy and Safety of the mRNA-1273 SARS-CoV-2 Vaccine. *N Engl J Med* 2021; **384**(5): 403-16.
